# A target HbA1c between 7 – 7.7% reduces macrovascular events in T2D regardless of duration of diabetes – a meta-analysis of randomized controlled trials

**DOI:** 10.1101/2021.03.11.21253387

**Authors:** Binayak Sinha, Samit Ghosal

## Abstract

**Introduction:** The target glycosylated haemoglobin (HbA1c) at which macrovascular benefits may be derived in type 2 diabetes (T2D) has never been clearly outlined. This meta-analysis was conducted on fifteen randomized controlled trials to highlight the association of HbA1c range with macrovascular events.

**Methods:** The association of different HbA1c clusters (intention to treat (ITT) and end-of-study [EOS]) range (less or equal than 6.5%, 6.6%–7.0%, 7.1%–7.7%) with macrovascular complications and also the combined effect of duration of T2D (< 10 years or ≥ 10 years) and HbA1c levels was assessed.

**Results:** Intensive glucose-lowering strategy resulted in a significant 13% reduction in non-fatal myocardial infarction (NFMI) (P=0.006). Based on HbA1c achieved, a significant 36% reduction in non-fatal stroke (P=0.008) and a 22% reduction in all-cause mortality (P=0.02) were observed in the group with HbA1c between 7.1% – 7.7% irrespective of diabetes duration. In the cohort, with diabetes duration <10 years, reduction of HbA1c in the range7.1% - 7.7% resulted in a significant 36% reduction in non-fatal stroke (NFS) (P<0.001).

**Conclusion:** This is probably the first meta-analysis highlighting the importance of treating patients with T2D to a target HbA1C of 7 – 7.7%, as this target is associated with reduction in macrovascular events.

## 1. Introduction

In the recent past multiple cardiovascular outcome trials (CVOT) in type 2 diabetes (T2D) have demonstrated an improvement of macrovascular outcomes with the usage of molecules like glucagon-like peptide receptor agonist (GLP1-RA) and sodium glucose co-transporter 2 inhibitor (SGLT2i). All these studies have attempted “glycaemic equipoise” in order to display the positive effects of the molecules and dispel any uncertainty that the benefits may be due to an improvement in glycaemia, although a meta-regression analysis of 12 CVOTs documented an average HBA1c reduction of 0.86%.(1) Multiple studies of glycaemic efficacy have been unable to prove that a reduction of HbA1C is convincingly associated with a reduction in adverse macrovascular outcomes, though an increase in HbA1c is associated with worsening macrovascular outcomes. (2,3) The significantly higher HBA1c levels in the placebo arm of the CVOTs as well as the HBA1c differential achieved therefore poses a serious challenge to interpretation of the outcomes data. (4) In addition, results from the CONTROL meta-analysis suggest a statistically significant 9% reduction in MACE associated with a 0.9% difference in HBA1c between the intensive and conventional glycaemic control arms. (5)

To further complicate the issue, the standard of care in all the CVOTs had set a target HBA1c of less than 7% or as per local guidelines or as per individual requirements based on advanced disease and complication status. Though this is in keeping with the ADA and AACE guideline of 2020, it is in deep contrast with the ACP 2018 guideline which recommends a HbA1c between 7%-8% for most patients. (6,7,8) The patient population recruited in the CVOTs with established CVD or with high risk of CVD, mean duration of diabetes more than 10 years and mean age around 60 years, would necessitate the individualization of HbA1c strategy setting the target between 7-8% as per the strategy of individualization. (9)

Thus, though a large amount of the benefit seen in the CVOT may be attributed to the singular pharmacology of GLP1-RA and SGLT2i, the beneficial effect of glycaemic control cannot be discounted. This meta-analysis was therefore conducted on the glycaemic efficacy studies, including end of study HbA1c, duration of diabetes and its impact on the macrovascular endpoints in these studies in an attempt to finally provide the correct glycaemic target to reduce macrovascular outcomes, irrespective of the molecule in use.

## 2.0. Method

This meta-analysis was conducted according to the recommendations of the PRISMA statement and registered with PROSPERO (ID: CRD42019122403).

### 2.1 Search strategy

A detailed literature search for relevant studies was conducted on the electronic biomedical databases Cochrane library, PubMed and Embase. The following keywords were used: MeSH terms: ‘Type 2 DM’; ‘cardiovascular diseases’. General terms: ‘non-fatal myocardial infarction’; ‘NFMI’; ‘non-fatal stroke’; ‘NFS’; ‘cardiovascular death’; ‘CV Death’; ‘all-cause mortality’; ‘hospitalisation for heart failure’; ‘hypoglycaemic agents’; ‘glycaemic control’; ‘tight glycaemic control’; ‘intensive glycaemic control’. The citations retrieved were screened according to pre-specified criteria (Fig. 1). Prospectively designed studies with an intensive arm and a control arm were chosen for the final analysis (n=15).

**Fig 1:**
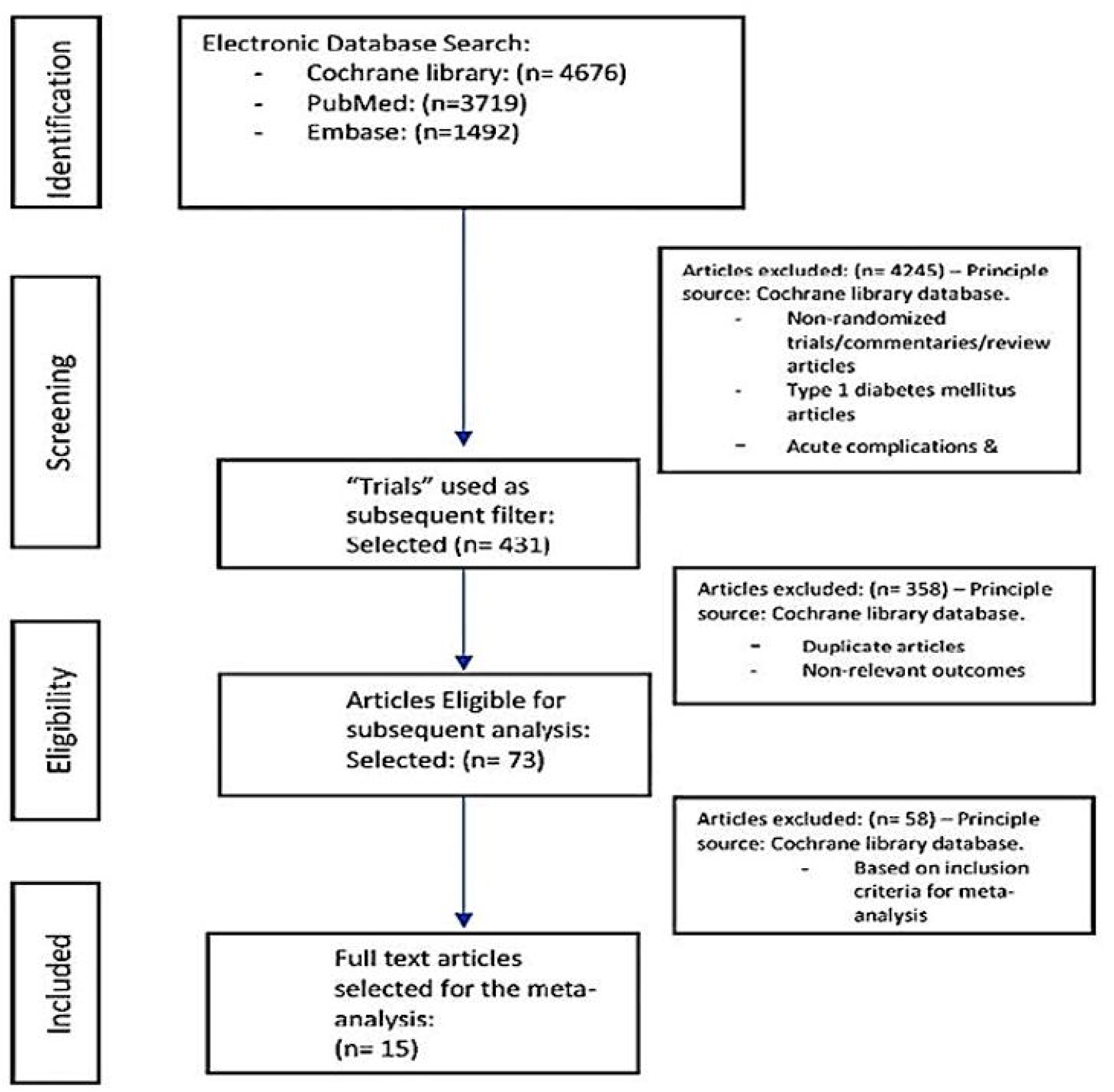
Study selection process.

An important distinction made in this analysis was selection of both the micro-and the macrovascular outcomes based on the current definitions (as used in recent CV outcomes trials). If the studies included in the present analysis had analysed the outcome measure of interest, but it did not conform to the modern definition of the term, it was not included in the meta-analysis.

### 2.2 Data extraction

Both authors independently conducted a web-based search for relevant citations dependent on the selected keywords. After identifying the citation from the web-based search, relevant data was extracted using the trial name, surname of the first author, year of publication, study population, place of origin of the study, design of the study, mean age, gender distribution, drugs in the intervention and control groups, dosages of agents in each group, background status related to cardiovascular disease, and duration of follow up. On identification of the basic database to work upon, further data extraction including the identification of NFMI, NFS, CV Death, ACM and hHF was undertaken. Additional filters included were, a cap on age above 18 years and clinical trials. No restrictions were placed based on language or date of publication. Any disagreements were resolved by conducting additional independent searches on a different day. After the initial process, a manual search was conducted jointly to identify the citations that met the inclusion criteria:

1. Randomized prospective trials on T2D
2. No cap on the number of patients recruited
3. Minimum duration of follow-up: 12 months
4. Reporting of the standardized outcome (macrovascular) end points in accordance with the accepted definitions as included in the CVOTs.
5. The control group included standard of care or placebo. The other baseline metabolic as well as CV risk parameters should also be matched. The process of data extraction is detailed in Fig. 1.

### 2.3 Quality assessment

The Cochrane risk of bias algorithm was used to assess quality of the studies. The assessment of the individual component of the Cochrane risk of bias algorithm was based on the attributes of those parameters detailed by Higgins and Altman^25^. The authors (based on mutual consensus) after reviewing the materials and methods section of all the selected citations, agreed that eight studies scored an unclear risk of blinding of outcome assessment because of insufficient information on random sequence generation and allocation concealment. One study (Home et al) showed bias due to blinding of outcome assessment, incomplete outcome data, and other biases. Issues related to unclear biases was also encountered in UDGP, UKPDS 33 & 34, Veterans Affairs, ACCORD, ADVANCE and VADT trials. (Supplementary fig 1) An additional web-based search was conducted to locate the original published protocol of the citations included in the analysis. Comparing the intended outcomes to be reported to the ones finally reported helped in identifying selective reporting and other biases, namely biases related to non-declaration of funding and conflict of interests, possibility of baseline imbalances which is difficult to decipher due to absence of publication of the trial protocol prior to conducting the trial, the degree of differences in the imbalances between the two comparative groups, and a pre-adjudicated and pre-specified hierarchical testing protocol.

### 2.4 Data synthesis and analysis

A detailed and up-to-date analysis from randomized prospective trials was conducted to assess the impact of intensive glycaemic control on NFMI, NFS, CV Death, ACM, and hHF in comparison to conventional therapy. Since the aim was to compare the two different strategies, we did not restrict the inclusion of studies to include the control group with placebo only. Having identified all the citations which reported the macrovascular outcomes of interest satisfying the pre-defined inclusion criteria, we went ahead with analysis.

Data analysis was conducted in a two-step manner:

Step 1: Analysis of the overall data (all 15 trials included) with an aim of identifying the impact of intensive glycaemic control versus conventional control on macrovascular outcomes based on the intension to treat strategy.

Step 2: A subgroup analysis was planned dividing the 15 included citations into two distinctive analytical strategy.

a. The impact of intensive glycaemic control versus conventional control on macrovascular outcomes based on the end-of-trial HBA1c i.e. based on the achieved HBA1c. The rationale to use this strategy was based on the ACP guidance which used the EOS HBA1c to proposed a relaxed HBA1c target for most T2D patients.
b. The impact of intensive glycaemic control versus conventional control on macrovascular outcomes based on the end-of-trial HBA1c and the duration of diabetes. Since a duration of diabetes of more than 10 years is considered as a high CV-risk factor, we took this cut-off as the parameter of interest.

A sensitivity analysis was planned for those parameters which demonstrated a statistically significant impact associated with a high degree of heterogeneity defined as a I^2^>75.

Data were analysed using the comprehensive meta-analysis software version 3 (Biostat Inc., Englewood, NJ, United States). Heterogeneity was assessed using the Cochrane Q and Higgins’s I^2^ test, and publication bias was assessed using funnel plots, with the precision (1/SE) plotted against the effect size. Individual effect size was assessed using hazard ratio (HR) with 95% confidence interval (CI). Effect size was assessed using the fixed or the random model depending on heterogeneity or on the possibility of the analysed study containing the true effect. Significant heterogeneity was defined as a P value <0.1 or a I^2^>75%.

## 3. Results

The meta-analysis was performed on a pooled population of 38,465 patients from fifteen citations, with 20,247 individuals in the intensive therapy arm and 18,218 individuals in the conventional treatment arm. However, UKPDS 34 was a sub study of UKPDS 33 and different outcomes were reported in the Veterans affairs and the VASCDM trials containing the same patient population. Hence, this meta-analysis was effectively performed on a pooled population of 37,559 individuals with 19,830 individuals in the intensive therapy arm and 17,728 individuals in the conventional treatment arm. The Cochrane risk of bias algorithm was used to assess quality of the studies included in the meta-analysis (Supplementary Fig. 1). In addition, publication bias was assessed using funnel plots of the individual endpoints.

The baseline characteristics of the studies included are presented in Table 1. The duration of the studies ranged from 2 to 10.7 years.

**Table 1.** Characteristics of studies included in the meta-analysis

| Studies | Country | Intensive arm (n) | Conventional arm(n) | Mean follow up (years) | Baseline HbA1c (mean/median) % | Expected HbA1c (Intensive arm) % | Achieved HbA1c-Mean (Intensive arm) % | Baseline DM duration (years) |
| --- | --- | --- | --- | --- | --- | --- | --- | --- |
| UGDP (1975) [10] | USA | 408 | 205 | 10 | NR | NR | NR | <1 |
| Veterans Affairs (1997) [11] | USA | 75 | 78 | 2.25 | 9.5 | <7.5 | 7.0 | 7.8 |
| UKPDS 33 (1998) [12] | UK | 3071 | 1138 | 10 | 7.1 | NR | 7.0 | <1 |
| UKPDS 34 (1998) [13] | UK | 342 | 411 | 10.7 | 7.3 | NR | 7.4 | <1 |
| VACS DM (1999) [14] | USA | 75 | 78 | 2 | 9.3 | <6.0 | <7.3 | 8.0 |
| KUMOMOTO (2000) [15] | Japan | 55 | 55 | 6 | 9.2 | <7.0 | 7.1 | 6.5 |
| PROactive (2005) [16] | Multinational | 2605 | 2633 | 2.9 | 7.9 | <6.5 | 7.0 | 8.0 |
| STENO-2 (2008) [17] | Denmark | 80 | 80 | 7.8 | 8.4 | <6.5 | 7.7 | 5.7 |
| ACCORD (2008) [18] | USA & Canada | 5128 | 5123 | 3.5 | 8.3 | <6.0 | 6.3 | 10.0 |
| ADVANCE (2008) [19] | Multinational | 5571 | 5569 | 5 | 7.5 | ≤6.5 | 6.5 | 8.0 |
| VADT (2009) [20] | USA | 892 | 899 | 5.6 | 9.4 | ≤6.0 | 7.0 | 11.5 |
| HOME (2009) [21] | Netherlands | 196 | 194 | 4.3 | 8.6 | 7.6 | 7.7 | 12 |
| RECORD (2009) [22] | UK | 321 | 323 | 5.5 | 7.9 | ≤7.0 | 7.5 | 7.9 |
| ADDITION EUROPE (2011) [23] | UK, Denmark & Netherlands | 157 | 161 | 5.3 | 7.0 | ≤7.0 | 6.6 | <1 |
| J-DOIT 3 (2017) [24] | Japan | 1271 | 1271 | 8.5 | 9.5 | <6.2 | 7.0 | 7.8 |

### 3.1 Impact of (positive/negative/neutral) intensive glucose-lowering strategy (irrespective of EOS achieved HbA1c) on macrovascular outcomes

Intensive glycaemic control resulted in a statistically significant 13% reduction in non-fatal myocardial infarction (NFMI) (95% CI: 0.78–0.96, P=0.006). There was no significant effect on nonfatal stroke (NFS) (HR: 0.84, 95% CI: 0.68–1.03, P=0.09), CV death (HR: 0.94, 95% CI: 0.84–1.06, P=0.38), all-cause mortality (HR: 0.98, 95% CI: 0.91–1.05, P=0.66) or hospitalisation for heart failure (hHF) (HR: 1.13, 95% CI: 0.88–1.44, P=0.32) (Fig 2).

**Fig 2:**
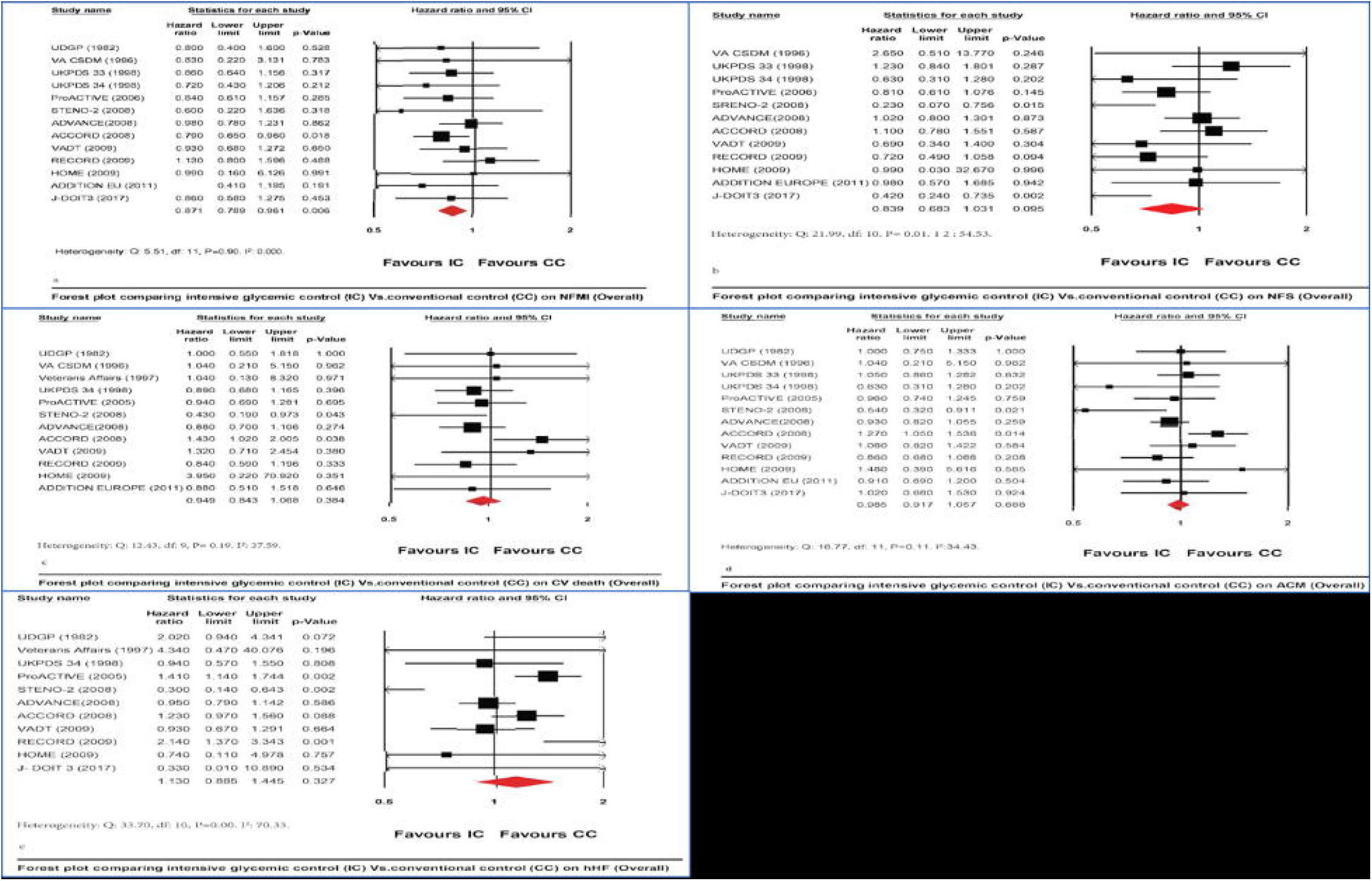
Impact of intensive glycaemic control versus conventional glycaemic control on macrovascular complications (Overall): A. Effect on non-fatal myocardial infarction: NFMI B. Effect on non-fatal stroke (NFS) C. Effect on cardiovascular death (CV Death) D. Effect on all-cause mortality E. Effect on hospitalization for heart failure (hHF)

### 3.2 Subgroup analysis and sensitivity analysis

In view of the fact, that, none of the outcomes of significance were associated with significant heterogeneity as per the pre-defined criteria, we went ahead with the two-step subgroup analysis. Step 1 involved splitting the EOS HBA1c as per the targets specified by most guidelines and step 2 used the criteria used in step 1 along with the duration of diabetes.

#### 3.2.1 Subgroup analysis based on EOS HBA1c

##### 3.2.1.1 End-of-study HbA1c ≤ 6.5%

There was a neutral impact on all the components of the macrovascular outcomes: NFMI (HR: 0.87, 95% CI: 0.70***–***1.07, P=0.20), NFS (HR: 1.04, 95% CI:0.85***–***1.27, P=0.65), CV death (HR: 1.10, 95% CI:0.68***–*** 1.77, P=0.68), all-cause mortality (HR:1.07, 95% CI: 0.79***–***1.46, P=0.63) and hHF (HR: 1.06, 95% CI:0.83***–***1.37, P=0.60) (Fig. 3).

**Fig 3:**
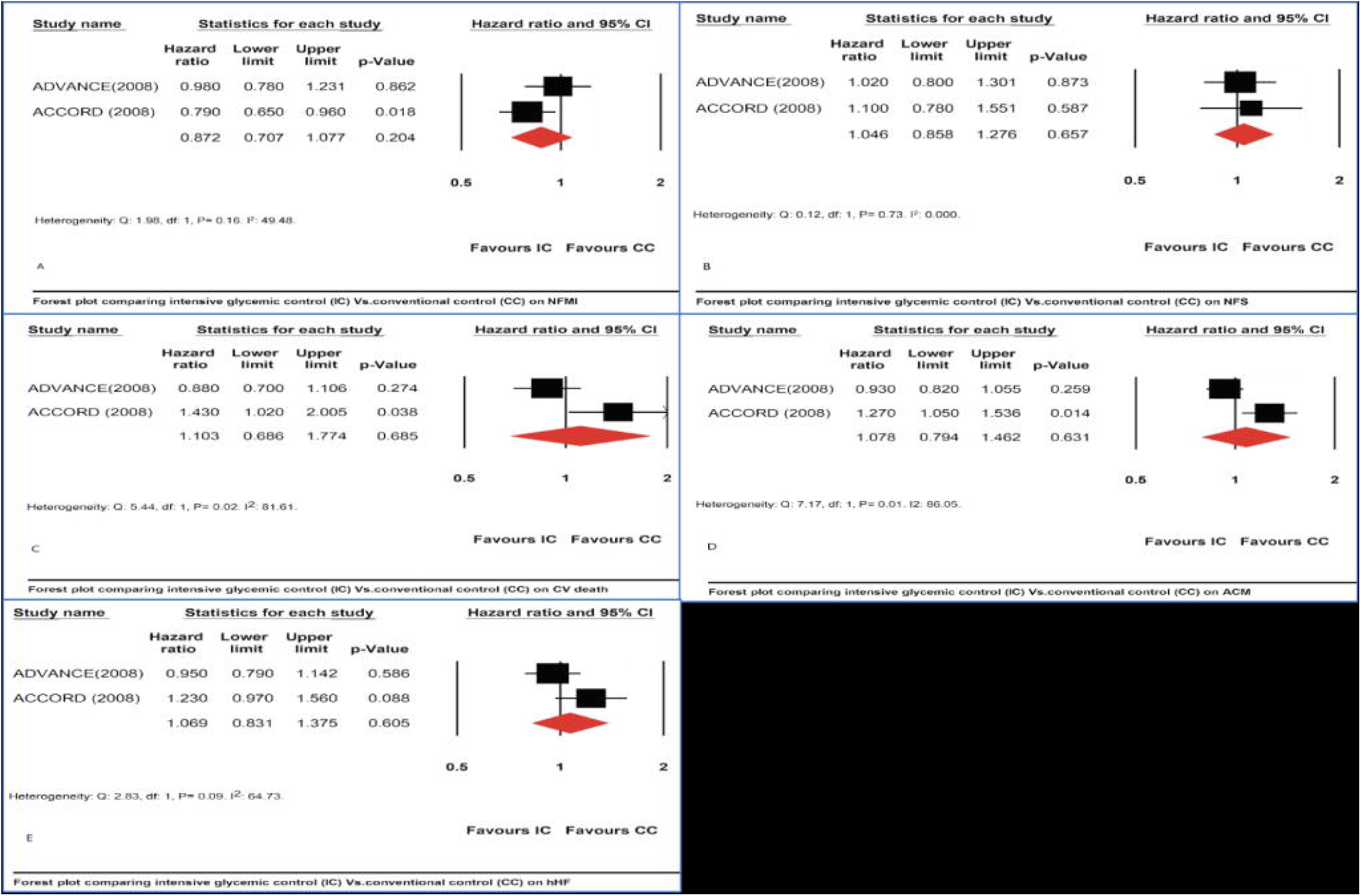
Impact of intensive glycaemic control versus conventional glycaemic control on macrovascular complications (EOS HBA1c□ 6.5%): A. Effect on non-fatal myocardial infarction: NFMI B. Effect on non-fatal stroke (NFS) C. Effect on cardiovascular death (CV Death) D. Effect on all-cause mortality E. Effect on hospitalization for heart failure (hHF)

##### 3.2.1.2 EOS HbA1c6.6%–7.0%

There was neutral effect of intensive glycaemic control on NFMI (HR:0.85, 95% CI:0.73***–***1.00, P=0.05), NFS (HR:0.84, 95% CI:0.69***–***1.10, P=0.07), CV death (HR:0.97, 95% CI:0.76***–***1.25, P=0.86), all-cause mortality (HR:1.00, 95% CI:0.89***–***1.13, P=0.90) and hHF (HR:1.15, 95% CI:0.78***–***1.68, P=0.46) (Fig. 4).

**Fig 4:**
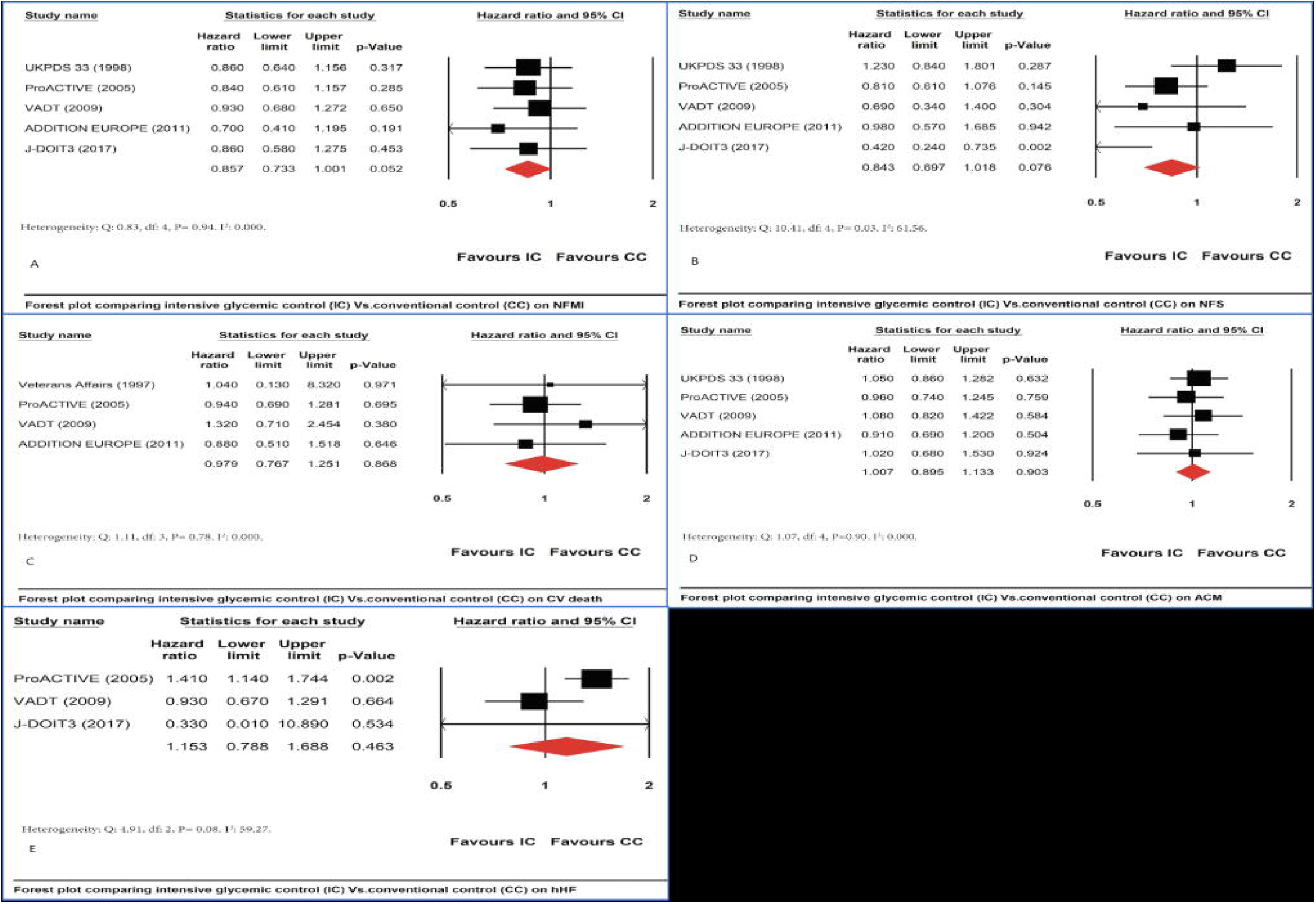
Impact of intensive glycaemic control versus conventional glycaemic control on macrovascular complications (EOS HBA1c 6.6%-7.0%): A. Effect on non-fatal myocardial infarction: NFMI B. Effect on non-fatal stroke (NFS) C. Effect on cardiovascular death (CV Death) D. Effect on all-cause mortality E. Effect on hospitalization for heart failure (hHF)

##### 3.2.1.3 EOS HbA1c7.1%–7.7%

There was a statistically significant 36% reduction in NFS (95% CI:0.46***–***0.89, P=0.008) and a statistically significant 22% reduction in all-cause mortality (95% CI:0.63***–***0.95, P=0.02). The effects on NFMI (HR:0.94, 95% CI:0.71***–***1.24, P=0.69), CV death (HR:0.83, 95% CI:0.67***–***1.02, P=0.08) and hHF (HR:0.88, 95% CI:0.32***–***2.41, P=0.80) were neutral (Fig. 5).

**Fig 5:**
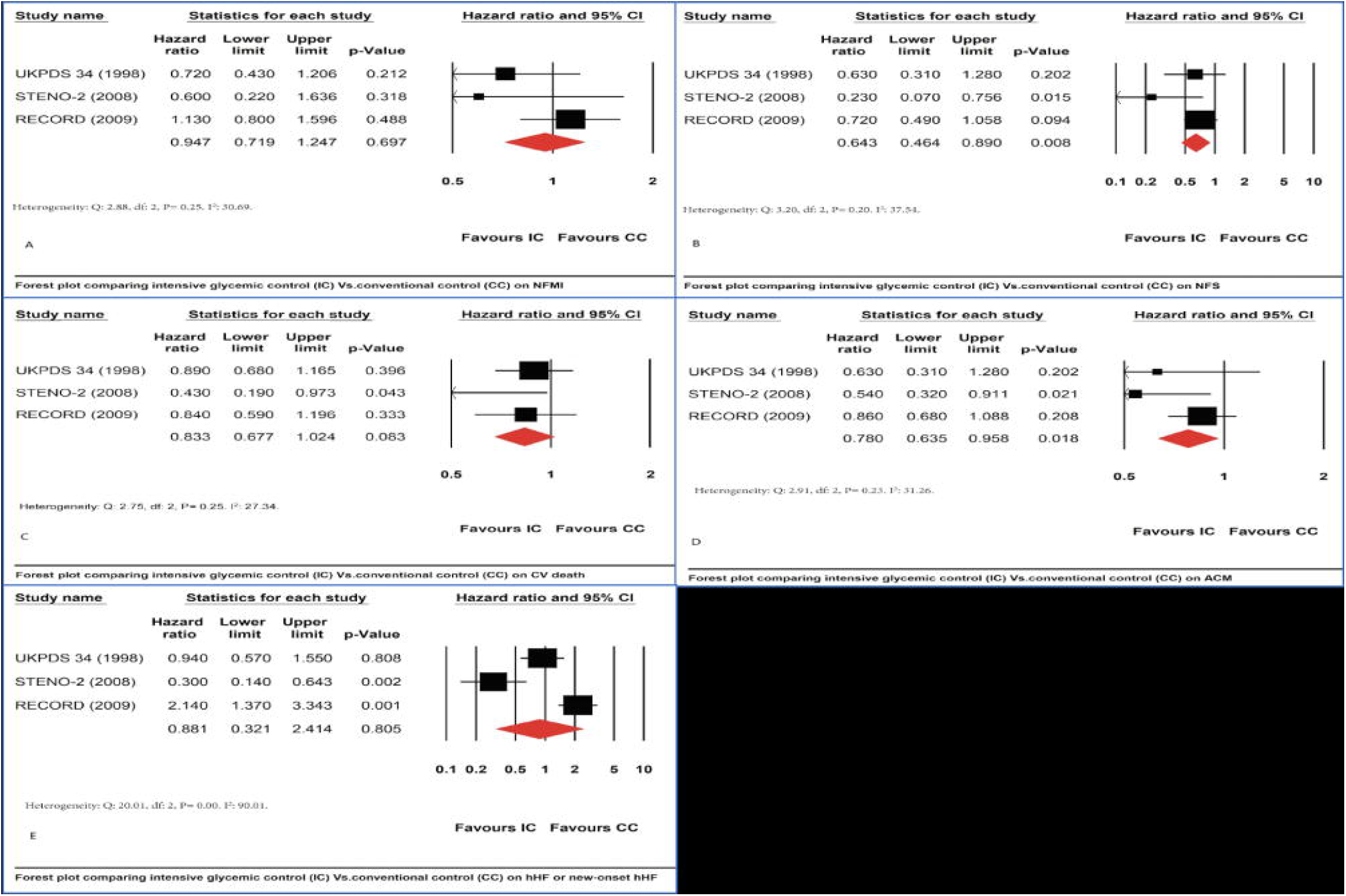
Impact of intensive glycaemic control versus conventional glycaemic control on macrovascular complications (EOS HBA1c >7.0%): A. Effect on non-fatal myocardial infarction: NFMI B. Effect on non-fatal stroke (NFS) C. Effect on cardiovascular death (CV Death) D. Effect on all-cause mortality E. Effect on hospitalization for heart failure (hHF)

#### 3.2.2 Subgroup analysis based on EOS HBA1c and duration of diabetes

The effects of intensive metabolic control on macrovascular complications was neutral based on the subgroup with a T2D duration <10 years and EOS HbA1c≤7.0%. (Supplementary Table 1).

There was a statistically significant 36% reduction in NFS (95% CI:0.46***–***0.89, P=0.008) achieved with intensive glycaemic control in patients with DM duration <10 years AND HbA1c 7.1%–7.7%. (Supplementary Table 1).

The combination of a T2D duration ≥10 years AND HbA1c 7.1%–7.7% was represented by a single trial and hence could not be analysed.

## 4. Discussion

### 4.1 Background

Arguably, the benefits of cardiovascular outcomes seen in the CVOTs are entirely due to the salutary effects of SGLT2i and GLP1RA in patients with T2D and not due to glycaemic control. Indeed, there have been studies indicating that a tight metabolic control may actually worsen cardiovascular outcomes in T2D. (18) However, there is little debate that a reduction of HbA1c is associated with an improvement in microvascular outcomes of diabetes. (25) In addition, the differing opinions of various august bodies like the ADA, AACE and the ACP have thrown up a conundrum as to which target of HbA1c should be targeted by treating physicians in order to protect the microvasculature and yet ensure that the macrovascular outcomes are not worsened. Multiple meta-analysis has tried to address this conundrum but have been restricted by their sample size, heterogeneity, data derived from type 1 and type 2 diabetes. Most importantly none of these meta analyses have categorized HbA1c levels into clusters (< 6.5%; 6.5 – 7%; 7 – 7.5% and > 7.5%), as has been done in this meta-analysis to pinpoint the exact target HbA1c range for maximal cardiovascular outcome benefit regardless of the molecule used to treat T2D. (26,27)

### 4.2 What this meta-analysis added

This meta-analysis was conducted on a large patient population (38,465 patients) including multiple trials (n=15) not included in the previous systematic reviews and meta-analyses. In addition to the intention-to-treat approach employed by most meta-analyses, this meta-analysis addresses the HbA1c as intention to treat (ITT) as well as the HBA1c achieved at end-of-study (EOS). It was felt sensible to evaluate end of study HbA1C since all the CVOT have presented their findings in the form of EOS between 6.8 and 7.7%. [7] Results from the ITT arm (target HBA1c range 6%-7.6%) indicated a significant reduction in NFMI (95% CI: 0.78–0.96, P=0.006). Subgroup analysis based on EOS HBA1c did not support targeting a HBA1c to either below 6.5% or below 7.0% in order to achieve macrovascular benefits. Achieving a HBA1c between 7%-7.7% seems to be the most appropriate target based on a 36% statistically significant reduction in NFS (95% CI:0.46***–***0.89, P=0.008) and a 22% reduction in ACM (95% CI:0.63***–***0.95, P=0.02). With a duration of diabetes less than 10 years, achieving HBA1c <7% was not associated with any macrovascular outcomes benefit, while a range between 7%-7.7% was associated with a robust 36% reduction in NFS (95% CI:0.46***–***0.89, P=0.008). In patients with T2D of more than 10 years, achieving a HBA1c below 7% was associated with a significant 17% reduction in NFMI (95% CI:0.70–0.97, P=0.02) associated with a 40% and 20% increased risk of CV death (95% CI:1.04–1.88, P=0.02) and all-cause mortality (95% CI:1.03–1.40, P=0.01), respectively. This worsening in CV death and ACM is entirely driven by the ACCORD study which is known to be skewed negatively towards good metabolic control.

### 4.3 Study limitations

Sample sizes differed between the HbA1c ranges studied in this analysis, since the available and eligible trials provided such a representative sample. Hence, any sub-group analysis would be biased towards such an imbalance. Data pertaining to a few end-points were reported by a single study; hence it was difficult to arrive at a definitive conclusion. We would like to include these areas as part of our research recommendations. Inclusion of other risk factors apart from target HbA1c and duration of diabetes may have provided different results. This was not done in our meta-analysis as we were restricted by only a limited number of prospective studies available for analysis. Any additional subgroups would have resulted in gross under-evaluation of the end-points.

### 4.4 Strengths of the meta-analyses

The biggest strength of this meta-analysis was the inclusion of large number of prospective studies. This allowed for analysis of one of the largest pool of data in comparison to most meta-analyses available till date. Another advantage was the inclusion of HbA1c achieved at EOS for analysis in contrast to the target HbA1c used in some meta-analyses. We predominantly used the random effect model, which is one of the most conservative modes of analysis, for estimation of the effect size. This helped minimise the risk of over-estimation of the effect.

### 5.0. Conclusion

This is probably the first meta-analysis to convincingly highlight that the maximum macrovascular benefits, by way of reduction of non-fatal MI, non-fatal stroke and all-cause mortality are achieved with a target HbA1C between 7 – 7.7%, whether the duration of T2D is less than or more than 10 years. This is in keeping with the ACP guidelines. Physicians should therefore aim for a target HbA1C of 7 – 7.7% regardless of which molecule is used for the treatment of T2D to improve the macrovascular outcomes of T2D.

## Supporting information

Supplementary figure 1: Quality of study assessment using the Cochrane Collaboration tool

## Data Availability

Data was available online.

## Author contributions

SG & BS conceptualised the idea and generated the hypothesis. SG & BS undertook the job of database search and study selection. SG conducted the meta-analysis. BS edited the whole document prior to submission.

## Abbreviations

ADA: American Diabetes Association
ACP: American College of Physicians
AACE: American Association of Clinical Endocrinologists
CDA: Canadian Diabetes Association
NICE: National Institute for Clinical Excellence
IDF: International Diabetes Federation
T2DM: Type 2 DM
DM: Diabetes Mellitus
CDC: The Centers for Disease Control and Prevention
HbA1c: Glycated haemoglobin C
CKD: Chronic Kidney Disease
GBD: Global Burden of Disease
CV deaths: Cardiovascular deaths
EOS: End-of-Study
ESRD: end-stage renal disease
HR: Hazard Ratio
CI: Confidence Interval
DPN: Diabetic Peripheral Neuropathy
IC: Intensive Control
CC: Conventional Control
hHF: Hospitalisation for Heart Failure
NFMI: Non-Fatal Myocardial Infarction
NFS: Non-Fatal Stroke
ACM: All-Cause Mortality

## Figure and table Legends

Supplementary figure 1: Quality of study assessment using the Cochrane Collaboration tool

Supplementary table 1: Impact of DM duration <10 years AND HbA1c<7.0%, DM duration <10 years AND HbA1c 7.1%–7.7%, and DM duration >10 years AND HbA1c<7.0% on NFMI, NFS, CV Death, ACM, and hHF.

