## Supplementary figures and images for "A target HbA1c between 7 – 7.7% reduces macrovascular events in T2D regardless of duration of diabetes – a meta-analysis of randomized controlled trials"

### Supplementary figure 1: Quality of study assessment using the Cochrane Collaboration tool

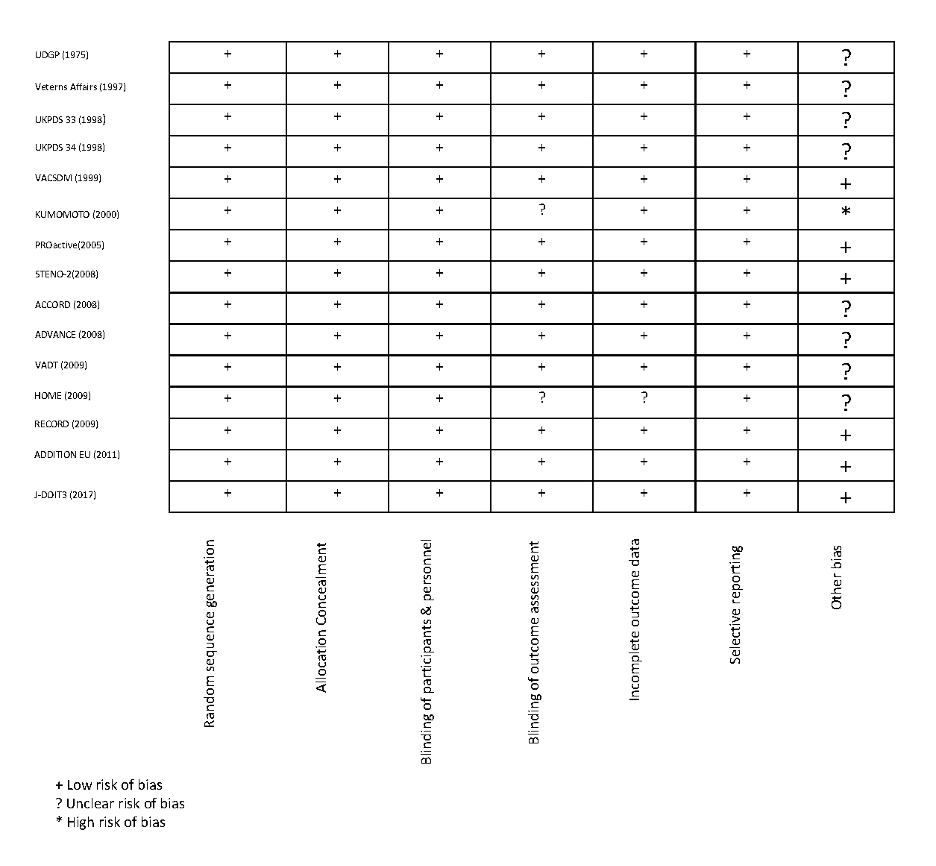
