## Supplemental Data 1 for "A target HbA1c between 7 – 7.7% reduces macrovascular events in T2D regardless of duration of diabetes – a meta-analysis of randomized controlled trials"

| DM duration <10 years AND HbA1c≤7.0% | HR | 95% CI | P-value | DF | Heterogeneity (I^2^) |
| --- | --- | --- | --- | --- | --- |
| Studies included for meta-analysis | UKPDS-33, PROactive, ADVANCE, ADDITION-EUROPE & JDOIT-3 | | | | |
| NFMI | 0.88 | 0.77-1.02 | 0.09 | 4 | 0.00 |
| NFS | 0.87 | 0.66-1.16 | 0.36 | 4 | 64.56 |
| CV Death | 0.90 | 0.75-1.07 | 0.23 | 4 | 0.00 |
| ACM | 0.95 | 0.87-1.05 | 0.36 | 4 | 0.00 |
| hHF | 1.17 | 0.81-1.71 | 0.39 | 4 | 64.26 |
| DM duration <10 years AND HbA1c 7.1%–7.7% | HR | 95% CI | P-value | DF | Heterogeneity (I^2^) |
| Studies included for meta-analysis | UKPDS-34, STENO-2 & RECORD | | | | |
| NFMI | 0.94 | 0.71-1.24 | 0.69 | 2 | 30.69 |
| NFS | 0.64 | 0.46-0.89 | 0.008 | 2 | 37.54 |
| CV Death | 0.83 | 0.67-1.02 | 0.08 | 2 | 27.34 |
| ACM | 0.85 | 0.64-1.13 | 0.28 | 2 | 66.57 |
| hHF | 1.15 | 0.84-1.56 | 0.36 | 2 | 90.01 |
| DM duration ≥10 years AND HbA1c≤7.0% | HR | 95% CI | P-value | DF | Heterogeneity (I^2^) |
| Studies included for meta-analysis | ACCORD & VADT | | | | |
| NFMI | 0.83 | 0.70-0.97 | 0.02 | 1 | 0.00 |
| NFS | 1.01 | 0.74-1.38 | 0.97 | 1 | 25.91 |
| CV Death | 1.40 | 1.04-1.89 | 0.03 | 1 | 0.00 |
| ACM | 1.21 | 1.03-1.41 | 0.02 | 1 | 0.00 |
| hHF | 1.12 | 0.92-1.35 | 0.26 | 1 | 45.42 |
